# CLINICAL AND DEMOGRAPHIC CHARACTERISTICS OF COVID-19 PATIENTS IN LAGOS, NIGERIA: A DESCRIPTIVE STUDY

**DOI:** 10.1101/2020.09.15.20195412

**Authors:** Ngozi Mirabel Otuonye, Testimony Jesupamilerin Olumade, Mercy Mayowa Ojetunde, Susan Abba Holdbrooke, Joy Boluwatife Ayoola, Itse Yusuf Nyam, Bamidele Iwalokun, Chika Onwuamah, Mabel Uwandu, Babatunde Lawal Salako, Akinola Abayomi, Akin Osibogun, Abimbola Bowale, Bodunrin Osikomaiya, Babafemi Thomas, Bamidele Mutiu, Nkiruka Nnonyelum Odunukwe

## Abstract

**Introduction:** COVID-19 is an emerging, rapidly evolving global situation, infecting over 25 million people and causing more than 850,000 deaths. Several signs and symptoms have been described to be characteristic of the disease. However, there is a dearth of report on the description of the clinical characteristics of the disease in patients from Nigeria. This study was designed to provide a description of the clinical and demographic characteristics of COVID-19 patients in Nigeria.

**Methods:** This study is a case series that includes patients that are evaluated between June and August 30, 2020, and diagnosed with COVID-19. Patient health records were reviewed and evaluated to describe the clinical characteristics on presentation.

**Results:** A total of 154 COVID-19 patients were included in this study, with a mean age (S.D.) of 46.16 (13.701). Most of the patients survived (mortality rate of 2.6%), and were symptomatic (89.6%). There were more males (74.7%) than females, and the most common symptoms were fever, breathing difficulty, dry cough and malaise. Co-morbidities were also present in almost half of the study participants (49.4%).

**Conclusion:** This study presents the most extensive description, to date, on the clinical and demographic characteristics of COVID-19 patients in Nigeria. Males are more likely than females to be infected with COVID-19 and the most occurring symptoms are fever, breathing difficulty, malaise, dry cough and chest pain. Old age and the presence of co-morbidities may also be associated with developing the severe disease.

## INTRODUCTION

COVID-19 is an emerging, rapidly evolving global situation which was first identified in December 2019, in the Hubei province of Wuhan, China. The disease that has grown into a pandemic is caused by the Severe Acute Respiratory Syndrome Coronavirus-2 (SARS-CoV-2) and has infected more than 25 million people globally, causing the death of more than 850,000 people [1,2]. The distribution of diseases is described in terms of person, place and time or who, where and when [3]. According to Chan *et al*. [4] and Andersen *et al*. [5], the initial outbreak seemed to be as a result of a zoonotic transmission from bats. However, as the outbreak continued, it was evident that human to human transmission of the virus via close contact was also possible through close contact with an infected person and through respiratory droplets, saliva or discharges from the nose when an infected person coughs or sneezes [6].

The clinical signs and symptoms of COVID-19 ranges in patients ranges from being absent (asymptomatic infections) to the mild or moderate infection with symptoms such as fever, dry cough, abdominal pain, fatigue, aches and pains, sore throat, breathing difficulty, diarrhoea, headache, conjunctivitis, loss of smell and/or taste, skin rash, and so on. Those who develop the severe disease exhibit more serious symptoms such as acute respiratory distress syndrome (ARDS), multiple organ failure and ultimately death [6-9]. Treatment remains largely supportive with the severe cases requiring oxygen supplementation and intensive care [10]. There are no approved vaccines yet for COVID-19.

The first case of COVID-19 in Nigeria was identified on 27th February 2020 in a visiting Italian. Since then, the epidemic in Nigeria has resulted in more than 54,000 cases and causing more than 1,000 deaths (mortality rate of 1.85%) as of September 1st, 2020 [11]. More than one-third (33.5%) of the cases in Nigeria were recorded in Lagos, Nigeria [11]. The epidemic in Nigeria has resulted in patients presenting with different symptoms and there is scarcity of information on the description of the clinical characteristics of COVID-19 patients in Nigeria. The only available study, to the best of our knowledge, describing the clinical characteristics of COVID-19 patients in Nigeria evaluated the first 32 COVID-19 patients in Nigeria [12]. This study was designed to provide a larger description of the clinical characteristics of patients presenting to an isolation centre in Lagos, Nigeria and confirmed to be infected with SARS-CoV-2 by real time Reverse Transcriptase Polymerase Chain Reaction (RT-PCR).

## METHODS

### Study Design and Participants

With ethical approval obtained from the Institutional Review Board (IRB) at the Nigerian Institute of Medical Research (NIMR), Yaba, Lagos, Nigeria, patient data were obtained and reviewed at the Mainland Infectious Disease Hospital, Yaba. Informed consent was also obtained from the study participants before their health records were obtained. This study is a case series that includes patients that are evaluated between June and August 30, 2020, and diagnosed with COVID-19. The IDIC is a central, comprehensive and integrated healthcare organization attending to COVID-19 patients in Lagos, the epicentre of the epidemic in Nigeria. All the patients included in this study were confirmed to have been infected with SARS-CoV-2 by a positive reverse transcriptase polymerase chain reaction test of nasopharyngeal, throat and blood samples. Clinical outcomes were also monitored and recorded.

### Data Collection and Statistical Analysis

Clinicians and trained research assistants reviewed patient health records retrospectively and copied them out to a standardized data collection form. Health records copied out include demographic information, signs and symptoms presented with, co-morbidities, and patient outcome. A formal sample size was not calculated for this study because the objective of the study was to describe the clinical characteristics of the patients who had enough information in their health records for analysis. Records were double entered into the forms before merging to reduce errors during data entry. Descriptive analyses were performed using Statistical Package for the Social Science (SPSS) version 25 (IBM, USA).

## RESULTS AND DISCUSSION

With COVID-19 being a novel disease, it is assumed that the immune system is naïve. Hence, the definition and description of the clinical characteristics after infection with the SARS-CoV-2 is important to foster early detection and control of the spread of the disease.

A total of 154 COVID-19 patients were included in this study. The mean age (Standard Deviation, [S.D]) of the study participants is 46.16 (13.701). Information about travel history was not collected because a nationwide lockdown was effected on 30^th^ March 2020 [13], and the study participants presented at the health centre between May and August, a time when community transmission of SARS-CoV-2 was already established in Nigeria [14]. Hence, it was assumed that the virus was contracted from other individuals infected with the virus.

Nearly all of the study participants (89.6%) were symptomatic (Table 1). This may explain why they presented at the health centre. This speaks to the health seeking behaviour of COVID-19 patients as being largely determined by the onset of symptoms. Asymptomatic patients are almost never aware that they are infected and, hence, do not need medical attention [15]. Four (2.6%) out of all the 154 patients included in this study died, giving a mortality rate of 2.6%. All four patients that died had co-morbidities – hypertension, diabetes, Lower Respiratory Tract Infection (LRTI) and Pneumonia that further complicated the disease.

**Table 1:** Baseline characteristics of COVID-19 patients at presentation N=154.

| Baseline Characteristics |  | N (%) |
| --- | --- | --- |
| Age: | Mean (S.D) | 46.16(13.701) |
|  | Range (Minimum – Maximum) | 16-83 |
| Gender: | Male | 115 (74.7) |
|  | Female | 39 (85) |
| Symptomatology: | Asymptomatic | 16 (10.4) |
|  | Symptomatic | 138 (89.6) |
| Patient Outcome: | Dead | 4 (2.6) |
|  | Survived | 150 (97.4) |

On admission, study participants presented with different symptoms (Table 2), with the most common ones being breathing difficulty (48.1%), fever (45.5%), dry cough (40.3%), malaise (48.7%), and chest pain (33.1%). About half of the study participants (49.4%) had pre-existing conditions, indicating co-morbidities at presentation with COVID-19, with the most occurring ones among the study participants being hypertension and Diabetes. From a meta-analysis consisting of 8697 patients with COVID-19 in China [16] and 3,062 patients with COVID-19 globally [17], the most occurring symptoms were fever, cough and malaise.

**Table 2:**
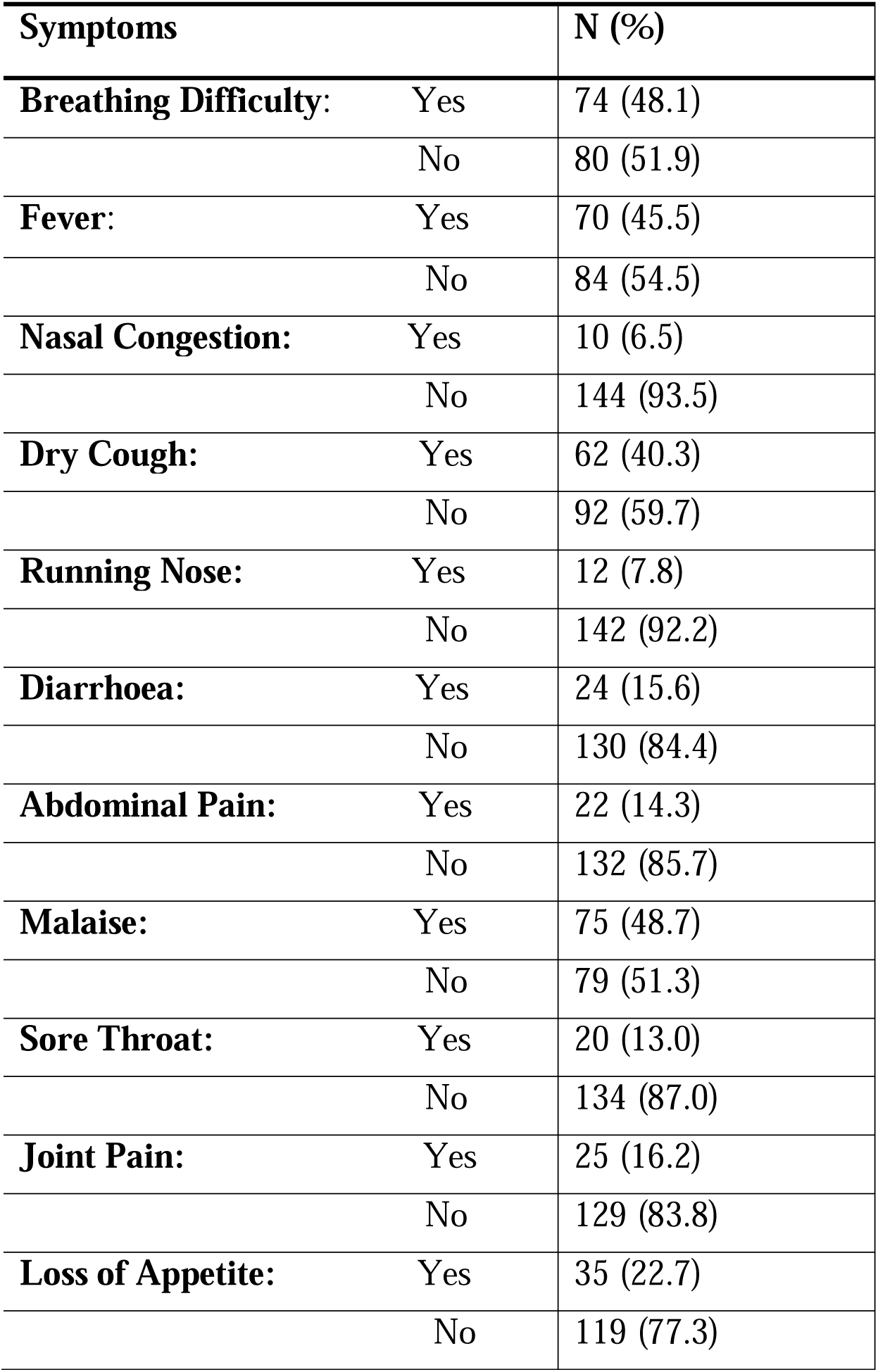
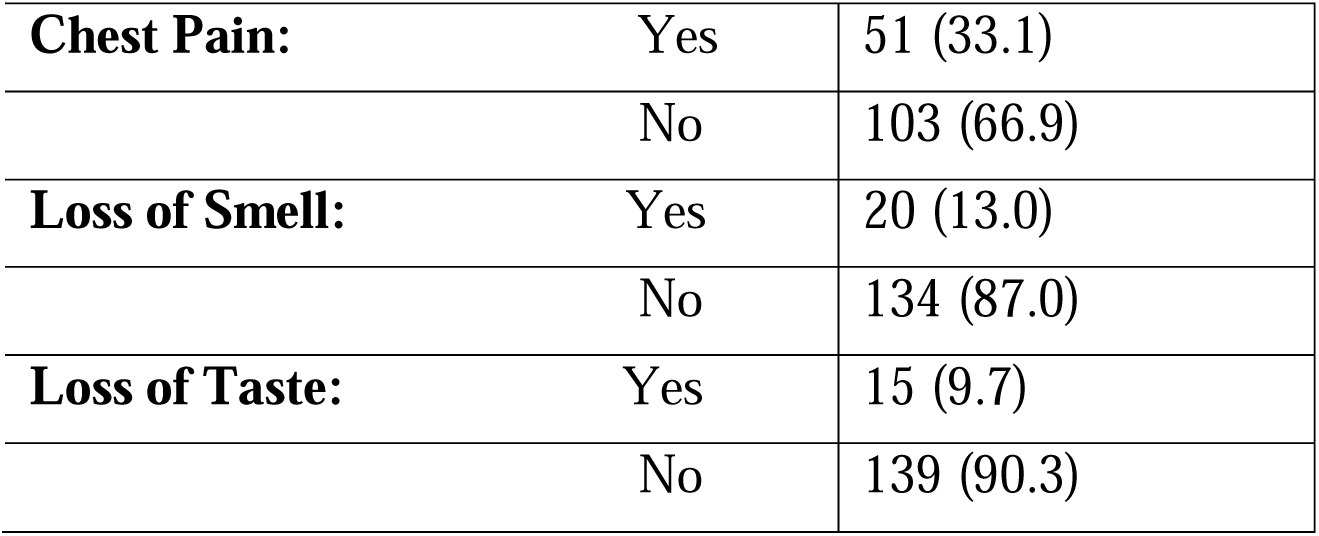
Symptoms of COVID-19 patients at presentation N=154.

| **Symptoms** |  | **N (%)** |
| --- | --- | --- |
| **Breathing Difficulty:** | Yes | 74 (48.1) |
|  | No | 80 (51.9) |
| **Fever:** | Yes | 70 (45.5) |
|  | No | 84 (54.5) |
| **Nasal Congestion:** | Yes | 10 (6.5) |
|  | No | 144 (93.5) |
| **Dry Cough:** | Yes | 62 (40.3) |
|  | No | 92 (59.7) |
| **Running Nose:** | Yes | 12 (7.8) |
|  | No | 142 (92.2) |
| **Diarrhoea:** | Yes | 24 (15.6) |
|  | No | 130 (84.4) |
| **Abdominal Pain:** | Yes | 22 (14.3) |
|  | No | 132 (85.7) |
| **Malaise:** | Yes | 75 (48.7) |
|  | No | 79 (51.3) |
| **Sore Throat:** | Yes | 20 (13.0) |
|  | No | 134 (87.0) |
| **Joint Pain:** | Yes | 25 (16.2) |
|  | No | 129 (83.8) |
| **Loss of Appetite:** | Yes | 35 (22.7) |
|  | No | 119 (77.3) |
| **Chest Pain:** | Yes | 51 (33.1) |
|  | No | 103 (66.9) |
| **Loss of Smell:** | Yes | 20 (13.0) |
|  | No | 134 (87.0) |
| **Loss of Taste:** | Yes | 15 (9.7) |
|  | No | 139 (90.3) |

**Table 3:**
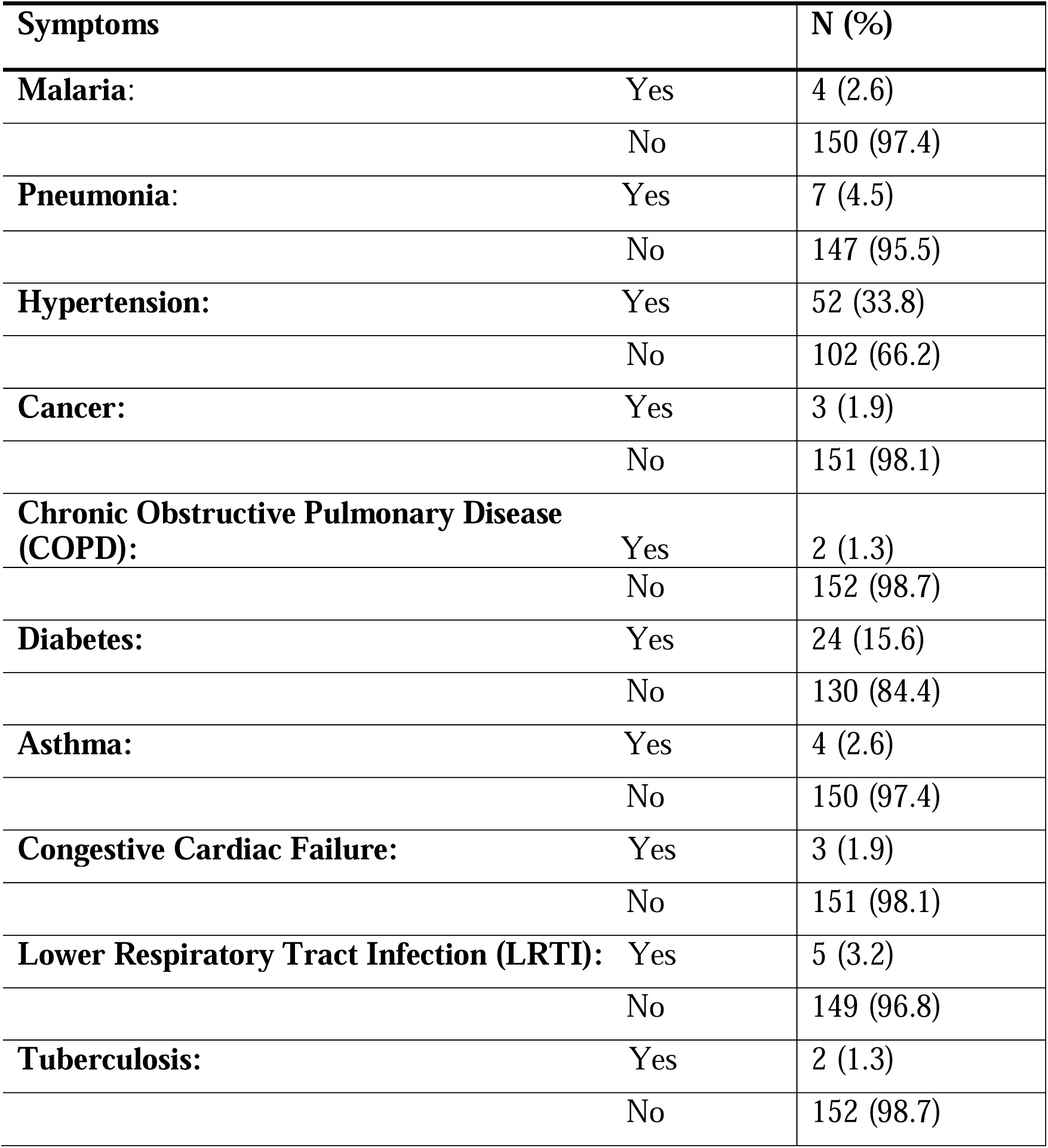
Co-morbidities in COVID-19 patients at presentation N=154.

| **Symptoms** |  | **N (%)** |
| --- | --- | --- |
| **Malaria:** | Yes | 4 (2.6) |
|  | No | 150 (97.4) |
| **Pneumonia:** | Yes | 7 (4.5) |
|  | No | 147 (95.5) |
| **Hypertension:** | Yes | 52 (33.8) |
|  | No | 102 (66.2) |
| **Cancer:** | Yes | 3 (1.9) |
|  | No | 151 (98.1) |
| **Chronic Obstructive Pulmonary Disease (COPD):** | Yes | 2 (1.3) |
|  | No | 152 (98.7) |
| **Diabetes:** | Yes | 24 (15.6) |
|  | No | 130 (84.4) |
| **Asthma:** | Yes | 4 (2.6) |
|  | No | 150 (97.4) |
| **Congestive Cardiac Failure:** | Yes | 3 (1.9) |
|  | No | 151 (98.1) |
| **Lower Respiratory Tract Infection (LRTI):** | Yes | 5 (3.2) |
|  | No | 149 (96.8) |
| **Tuberculosis:** | Yes | 2 (1.3) |
|  | No | 152 (98.7) |

None of the patients needed ventilator-assisted breathing, developed ARDS, or had organ failure. The disparity between the severity of the disease in Nigeria (and by extension the African continent) and the rest of the world has been discussed by other studies with some reasons given for the observed disproportionality in outcomes include the relatively young population in Africa, lower rates of obesity, poor access to health care, familiarity of the continent with outbreaks of infectious diseases, impact of health disparities, and lower socioeconomic factors in Africa [18-20]. Although, on the other hand, reports from other continents show higher morbidity and mortality in people of African heritage [21, 22].

About three-quarter of the study participants were men, a trend also observed in other studies [7,16]. SARS-CoV and MERS-CoV have also been reported to infect more males than females, a phenomenon that could be attributed to protection from the X chromosome and sex hormones, known to play significant roles in innate and adaptive immunity [23, 24]. Other possible reasons proposed for the increased susceptibility of men to COVID-19 include biological reasons such as a higher expression of angiotensin-converting enzyme (ACE 2, a reception coronaviruses) in males than females; and behavioural reasons – higher levels of smoking and drinking in men, and irresponsible attitude, reviewed by [25] The results of this study suggest that older adults are more likely to be infected with SARS-CoV-2 and be more susceptible to develop the severe disease, a trend that may be due to the reduced immunity in older adults. This trend is well established globally, reviewed by [26]. In this study, the four patients who died were all older than 60 years. It has also been reported that old age, obesity and the presence of co-morbidities may be associated with increased mortality [27].

The results of this study have implications for public health, particularly the surveillance and diagnosis of SARS-CoV-2. The clinical signs and symptoms may not be specific only to COVID-19, public health workers and clinicians should consider these symptoms in addition to epidemiological information such as travel history, contact with suspected or confirmed cases of COVID-19 within 14 days, and the presence of co-morbidities. In addition, although only 16 (10.4%) of the study participants were asymptomatic, it is known that they can be infective [28], and they should be identified as quickly as possible through the sampling of the contacts of each confirmed case, so that the spread of the virus can be controlled. Furthermore, this study highlights the importance of the integration of different intervention measures to contain the spread of the virus – highly effective contact tracing, isolation or quarantine, social distancing, the use of hand sanitizers, wearing of masks, and washing of hands [29].

This study has its limitations. First, the study is limited by the sample size. Only 154 patients were included in the study. Suspected, but not confirmed, cases were not included in this study. More patient data from other health centres around the country should be added to obtain a more comprehensive understanding of the clinical characteristics of the disease in the country. Second, patient data such as travel history was also incomplete at the time this study was conducted, hence the assumption of local transmission. However, this study presents, to the best of our knowledge, the largest report describing the clinical characteristics of COVID-19 patients in Nigeria.

In conclusion, this study presents the most extensive description, to date, on the clinical and demographic characteristics of COVID-19 patients in Nigeria. From the results of this study, males are more likely to be infected with COVID-19, and the most occurring symptoms are fever, breathing difficulty, malaise, dry cough and chest pain. Old age and the presence of co-morbidities may also be associated with disease severity.

### Summary Table

- The only study from Nigeria on the subject so far (32 patients) reports fever and dry cough as the most common symptoms.
- We present data from 154 COVID-19 patients, with the most common symptoms being fever, breathing difficulty, malaise, dry cough and chest pain.
- Males are more likely to be infected; old age and co-morbidities are associated with disease severity.

### Summary Sentence

This work represents an advance in biomedical science because we present the most extensive description so far on the clinical and demographic characteristics associated with COVID-19 in Nigeria.

### Ethical Approval and Consent to Participate

Approval to conduct this study was received from the Institutional Review Board (IRB) at NIMR, Yaba, Lagos. Informed consent was obtained from the participants before they were enrolled in the study.

### Consent for Publishing

All authors gave their consent for publishing

### Availability of Supporting Data

Besides those presented in the study, there are no other supporting data for this manuscript.

## Data Availability

Besides those presented in the study, there are no other supporting data for this manuscript.

## Competing Interests

All authors declare no competing interests.

## Funding

The PI and co-authors financed this study

## Authors’ contributions

NMO conceptualized and designed the study

TJO collated, entered, analysed and interpreted data

MMO, SAH, JBA, IYN collected data at the isolation site

TJO and MMO wrote the initial draft of the manuscript

MU provided some consumables used for the project

BI, CO, NNO reviewed the proposal

BLS contributed some funds for the project

AA, AB, AO, BO, BT, BM are staffs of the isolation centre and they provided the patients’ data being analysed

All authors approved the final version of the manuscript.

## Acknowledgments

The authors wish to thank the members of staff of the Nigerian Institute of Medical Research (NIMR), Yaba, and the Mainland Infectious Disease Hospital, Yaba, Lagos.

## Notes

### Competing Interest Statement

The authors have declared no competing interest.

### Author Declarations

Nigerian Institute of Medical Research (NIMR) Institutional Review Board (IRB), Yaba, Lagos, Nigeria.

